# Twelve-month follow-up of immunogenicity and safety of fractional and standard booster doses of the Pfizer-BioNTech COVID-19 vaccine in adults primed with ChAdOx1, BBIBP-CorV, or GAM-CoV-Vac: a randomised controlled trial

**DOI:** 10.1101/2025.08.29.25334703

**Authors:** Tsetsegsaikhan Batmunkh, Eleanor F G Neal, Otgonjargal Amraa, Nadia Mazarakis, Bolor Altangerel, Naranbaatar Avaa, Lkhagvagaram Batbayar, Khishigjargal Batsukh, Kathryn Bright, Tsogjargal Burentogtokh, Lien Anh Ha Do, Gantuya Dorj, John D Hart, Otgonbold Jamiyandorj, Khulan Javkhlantugs, Sarantsetseg Jigjidsuren, Frances Justice, Shuo Li, Khaliunaa Mashbaatar, Kerryn A Moore, Narantuya Namjil, Cattram Duong Nguyen, Batbayar Ochirbat, Unursaikhan Surenjav, Helen Thomson, Bilegtsaikhan Tsolmon, Paul V Licciardi, Claire von Mollendorf, Kim Mulholland

## Abstract

**Background:** COVID-19 vaccine booster doses counteract waning immunity and vaccine escape by emerging variants. We evaluated long-term immunogenicity and safety of fractional and standard BNT162b2 vaccine booster doses in Mongolia.

**Methods:** In this randomised, controlled, non-inferiority trial, adults primed with two doses of ChAdOx1-S, BBIBP-CorV, or Gam-COVID-Vac were randomised (1:1) to receive a 15μg (fractional dose) or 30μg (standard-dose) BNT162b2 booster. Geometric mean ratios (GMR) of IgG and surrogate virus neutralising test (sVNT) levels (Wuhan-Hu-1 and Omicron B.1.1.529) were compared over 12 months. SARS-CoV-2 infections, adverse and serious adverse events (SAEs) were documented. ClinicalTrials.gov Identifier: NCT05265065.

**Results:** Of 601 participants randomised between May 27th and September 30th, 2022, 2 (0.3%) were lost to follow-up and 19 (3.2%) withdrew by 12 months. IgG levels declined from 28 days to six months, stabilising thereafter. At 12 months, IgG levels were lower in the fractional compared with standard arm for ChAdOx1-S primed participants (GMR 0.78 [95% CI 0.63–0.96], *p*=0.017). At six and 12 months, the median sVNT inhibition percentages were comparable by study arm and priming strata. Documented SARS-CoV-2 infections occurred in 25 participants (fractional dose arm n=12; standard dose arm n=13). From 28 days, 228 undocumented infections (≥ 1.2-fold IgG increase) occurred (fractional arms n=112; standard arm n=116). SAEs (n=41) were balanced between arms, with no severe vaccine-related AEs and SAEs were reported.

**Conclusions:** Fractional and standard dose BNT162b2 boosters demonstrated comparable immunogenicity and favourable safety. Fractional COVID-19 vaccine booster doses may improve vaccine acceptability due to lower reactogenicity.

## Introduction

The COVID-19 pandemic has driven global efforts to vaccinate populations to control the spread of SARS-CoV-2 and mitigate severe disease outcomes. Despite the success of initial vaccination campaigns, the emergence of new variants and waning immunity have underscored the necessity for booster doses to sustain protective immunity. Optimising booster dose strategies is critical, particularly in resource-limited settings, to maximise vaccine impact and ensure equitable access. Fractional dosing, with lower booster doses, has been proposed to balance immunogenicity and reactogenicity while reducing costs, extending vaccine supplies, and increasing acceptance.[1]

In our previous study, we demonstrated the short-term immunogenicity and safety of a fractional dose BNT162b2 (Pfizer-BioNTech) booster relative to the standard formulation in adults primed with ChAdOx1-S (Oxford-AstraZeneca), BBIBP-CorV (Sinopharm), or Gam-COVID-Vac (Gamaleya).[2] These findings are consistent with other studies suggesting fractional mRNA doses may achieve similar immunogenicity with fewer adverse events than standard doses.[3, 4] A 2021 double-blinded randomised controlled trial (RCT) in Thailand found that a half-dose ChAdOx1-S booster following two CoronaVac doses provided non-inferior immunogenicity to a full-dose booster, with lower rates of fever and myalgia at 14 and 90 days post-boost.[3] Similarly, a sub-study of the UK’s COV-BOOST phase 2 RCT evaluated a fourth booster with either a full-dose BNT162b2 (30 μg, Pfizer-BioNTech) or a half dose of mRNA-1273 (50 μg, Moderna).[4] While the half-dose mRNA1273 booster had higher mRNA content than the full-dose BNT162b2, it demonstrated superior immunogenicity at day 14 and was well tolerated.[4] However, longitudinal data following fractional dosing, particularly in low- and middle-income countries (LMICs), remains a critical gap in evidence.

Understanding the durability of the immune response and the sustained safety profile over an extended period is crucial for informing public health strategies. This study aims to assess and compare the immune response, measured as binding antibodies (IgG ELISA) at six and 12 months following standard versus fractional doses of the BNT162b2 vaccine given as a booster dose for three priming strata. This study also evaluated the safety of the booster dose regimen over three months. It provides valuable insights into the long-term booster performance in populations primed with different COVID-19 vaccines widely used in low- and middle-income countries.

## Materials and methods

### Trial design

The design of this double-blind, controlled trial has been described previously.[2] Briefly, this phase 3, randomised controlled non-inferiority trial in Mongolia compares the immunogenicity and safety of a fractional (15μg) vs standard dose (30μg) BNT162b2 COVID-19 booster, given via intramuscular injection, in adults primed with two doses of ChAdOx1-S, BBIBP-CorV, or Gam-COVID-Vac.

### Study vaccine, storage, and administration

BNT162b2 is a lipid nanoparticle-formulated, nucleoside-modified mRNA vaccine that encodes trimerised SARS-CoV-2 spike glycoprotein.[5, 6] BNT162b2 encodes the SARS-CoV-2 full-length spike, modified by two proline mutations to lock it in the prefusion conformation and more closely mimic the intact virus with which the elicited virus-neutralizing antibodies must interact.[5-7] mRNA vaccines use the pathogen’s genetic code as the vaccine; this then exploits the host cells to translate the code and then make the target spike protein.[5] The protein then acts as an intracellular antigen to stimulate the immune response.[5] The vaccine RNA is formulated in lipid nanoparticles for more efficient delivery into cells after intramuscular injection.[8]

The study vaccine was stored at the central vaccine storage facility at the National Centre for Communicable Diseases (NCCD), following the manufacturer’s recommendations, at -70°C ±10°C (shelf life of six months). Once thawed, it was stored at 2–8°C for up to 31 days. The study vaccine lot number was FFJ1966.

Booster doses of BNT162b2 were administered via intramuscular injection in the middle of the deltoid, above the deltoid tuberosity, using a 25 mm needle. No concomitant vaccines were administered. The study vaccine was stored at the central vaccine storage facility.

### Participants, randomisation, and blinding

Eligible participants were healthy adults aged 18 years or older who had previously received two doses of BBIBP-CorV, ChAdOx1-S, or Gam-COVID-Vac vaccines. Enrolled participants were randomly assigned (1:1), stratified by age (18 - <50 and ≥ 50 years), and priming vaccine to receive 15 µg (fractional dose) or 30 µg (standard dose) of the BNT162b2 vaccine, and followed for 12 months. Participants were followed for two years, with blood draws pre-boost and at 28 days, six months, 12 months, 18, and 24 months post-boost. Study staff administering the vaccine were unblinded. Participants and the clinical study team were blinded until the 28-day visit. Laboratory staff and statisticians were blinded to treatment allocation during specimen analysis and statistical planning.

### Procedures

Blood samples were collected from participants at baseline, 28 days, six- and 12-months post-booster to measure antibody levels. Blood samples were processed for serum and stored at −80°C. Anti-spike IgG was measured using SARS-CoV-2 QuantiVac S1 IgG ELISA kits (EUROIMMUN Medizinische Labordiagnostika AG, Lübeck, Germany). Results were reported as relative units (RU)/mL and then converted to binding antibody units (BAU)/mL per the manufacturer’s instructions (multiplication factor = 3.2). Results <25.6 BAU/mL were considered negative, 25.6 - 35.2 BAU/mL were considered borderline, and results >35.2 BAU/mL were considered positive. The percentage inhibition of the interaction between SARS-CoV-2 receptor binding domain (RBD) (Wuhan-Hu-1 or Omicron B.1.1.529 (BA.1)) and ACE2 by neutralising antibodies was measured using the C-PASS surrogate virus neutralising test (sVNT) (GenScript cPass SARS-CoV-2, New Jersey, USA). Results were reported as a percentage (%) of inhibition calculated from the formula; % inhibition = [1 - (optical density value of sample/optical density value of the background)] x 100%. Results < 30% inhibition are considered negative, while ≥ 30% inhibition is considered positive for surrogate neutralising antibodies.

The safety of the booster dose regimen was evaluated by monitoring adverse events (AEs) and serious adverse events (SAEs). Solicited AEs were collected for the first three months post-booster, and unsolicited AEs were recorded as they occurred. SAEs were tracked continuously, with all events documented in REDCap, including severity (mild, moderate, severe, potentially life-threatening, or fatal), outcome (resolved, resolved with sequelae, ongoing, fatal, unknown) and relationship to the intervention (unrelated, possible, probably, definite), with a MedDRA classification assigned.

SARS-CoV-2 infections were monitored and documented throughout the study period. Participants were asked if they had tested positive for SARS-CoV-2 by rapid antigen test (RAT) or polymerase chain reaction (PCR) at time of enrolment and at each follow-up visit. If participants had tested positive, study staff recorded details including the date of the positive test result, whether the participant had symptoms consistent with COVID-19, and the clinical spectrum of the illness (mild, moderate, severe, critical). From March 2023, participants received monthly SMS reminders between the six- and 12-month visits, asking them to contact the clinic to provide samples for more detailed genotyping using next-generation sequencing if they had COVID-19 symptoms or had a confirmed SARS-CoV-2 infection. Additionally, a blood test was arranged for 28 days after the initial positive test result.

### Statistical analysis

#### Participant recruitment and follow-up

The methods for summarising the baseline characteristics for this study population have been reported in the previous publication.[2] Here we add the summary of the history of SARS-CoV-2 exposure before boosting and participant retention, including attendance, withdrawals, and loss to follow-up at each study visit up to 12 months.

#### Assessment of Missingness

To evaluate data completeness across key variables, we systematically assessed missingness for immunogenicity, descriptive, and denominator data. This included calculating missing data rates for anti-spike IgG levels, sVNT inhibition data (Wuhan-Hu-1 and Omicron BA.1), and baseline characteristics within each analysis population. Missingness patterns and associations with study variables were evaluated using cross-tabulations and Chi-squared tests, stratified by study arm and priming strata.

Analysis populations were defined in advance, as all randomised participants who received a study vaccine, according to the vaccine received, documenting any initial exclusions due to missing baseline or follow-up data. Missing data rates were calculated for the full enrolled cohort and within each analysis population to clarify any differences due to prior exclusions.

#### Estimand framework and analysis approach

To ensure the statistical analysis met the study objectives, we used the Estimand Framework.[9] Estimand-to-analysis tables, which relate each of the five estimand attributes to the analytical methods, were specified in the statistical analysis plan for each endpoint.[2] The estimands for the long-term immunogenicity endpoints were the geometric mean ratio (GMR) of IgG and median surrogate virus neutralisation levels at six and 12 months after boosting with a single fractional (15 μg) dose compared to a single standard (30 μg) dose of BNT, in healthy people aged ≥18 years in Mongolia, regardless of breakthrough SARS-CoV-2 infections or receipt of a 4^th^ dose (self-initiated). SARS-CoV-2 infection post-boosting was treated as a post-randomisation (intercurrent) event using the Treatment Policy strategy.[9] Supplementary analyses using the Hypothetical Strategy for infections occurring >14 days post-boost were planned but not conducted due to the small number of events (n=25). Additionally, the balanced case distribution between study arms and similar vaccine performance observed in the Day-28 analysis, made differential effects by study arm unlikely even with potentially low ascertainment.[9] Missing data were managed through complete case analysis.

#### Long-term anti-spike IgG levels

To assess long-term immunogenicity, GMRs were estimated by fitting linear regression models to the log-transformed anti-spike IgG levels, adjusted for age group, priming strata, study day of blood draw, duration between 1st and 2nd, and 2nd and 3rd doses, and baseline anti-spike IgG levels. The primary endpoint (binary seroresponse at 28 days) was analysed using a non-inferiority framework, whereas the six- and 12-month immunological data have been analysed as continuous variables using a superiority framework, with results interpreted in terms of effects sizes of GMRs and 95% confidence intervals (95% CI). GMRs were calculated by taking the antilogarithm of the mean difference from these models, and an interaction term between study arm and priming strata was included to estimate GMRs within each priming stratum. By visit, IgG levels were plotted by study arm and priming strata, with median and interquartile ranges (IQR) to highlight trends and variability across time points. We tabulated any increase in anti-spike IgG levels since the last visit. Differences in the percent of participants with increased binding antibody titres between study arms and by priming strata were assessed using the Chi-squared test, with α set to < 0.05.

#### Long-term sVNT inhibition responses to Wuhan-Hu-1 and Omicron BA.1

To evaluate and compare the durability of neutralising responses over time, the median and IQR for sVNT percent inhibition against Wuhan-Hu-1 and Omicron BA.1 were calculated at each time point, stratified by study arm and priming strata. Results are presented numerically and graphically.

#### Documented and undocumented intercurrent SARS-CoV-2 infections

To identify SARS-CoV-2 infections over the study period, an epidemiological curve was generated, summarising documented intercurrent SARS-CoV-2 infections by study week. To review antibody response data and identify potential undocumented intercurrent infections, we generated spaghetti plots stratified by study arm and priming strata arm. Each line represents an individual participant’s spike-IgG titres across time points (baseline, 28 days, six months, and 12 months), highlighting antibody trajectories over time. These plots allowed us to visually assess the direction and magnitude of changes between visits, offering insights into potential instances of undetected infections.

To identify potential undocumented intercurrent SARS-CoV-2 infections, we applied a fold change threshold of ≥1.2 in anti-spike IgG levels between visits (Day 28 to six months and six to 12 months). Participants meeting this threshold without a reported SARS-CoV-2 infection during these periods were classified as having a potential intercurrent infection.

We tabulated documented SARS-CoV-2 infections and fold changes of ≥1.2 in anti-spike levels without a documented infection (indicative of unreported intercurrent infection). Differences in the incidence of documented and undocumented infections between study arms and by priming strata were assessed using the Chi-squared test, with α set to < 0.05.

#### Adverse and serious adverse events

AEs and SAEs were tabulated by severity, causality, and MedDRA System Organ Class (SOC). Events outside the 12-month window (defined as ≥ 335 to ≤ 395 days post-randomisation) were excluded. Descriptive statistics, including counts and percentages, were provided, with counts of SAEs by randomisation group compared using the Chi-squared test, with α set to < 0.05.

All statistical analyses were conducted using Stata version 18.1.

#### Ethics

The Mongolian Ethics Committee of the Ministry of Health (Decision #273, 05 April 2022) and the Royal Children’s Hospital Human Research Ethics Committee (HREC/81800/RCHM-2021) approved the trial. The trial was conducted per the Declaration of Helsinki and Good Clinical Practice guidelines. All participants provided written informed consent before enrolment. The trial was registered with ClinicalTrials.gov (identifier NCT05265065). The study protocol has been published previously.[2]

## Results

### Participant recruitment and follow-up

The baseline characteristics of participants in this trial have been described previously (Supplementary Table 1).[2] At six months, 290 fractional dose and 285 standard dose participants were analysed (Figure 1). At 12 months, 287 fractional dose and 285 standard dose participants were analysed. No participant reported receipt of a 4^th^ dose of a COVID-19 vaccine in the community.

**Figure 1.**
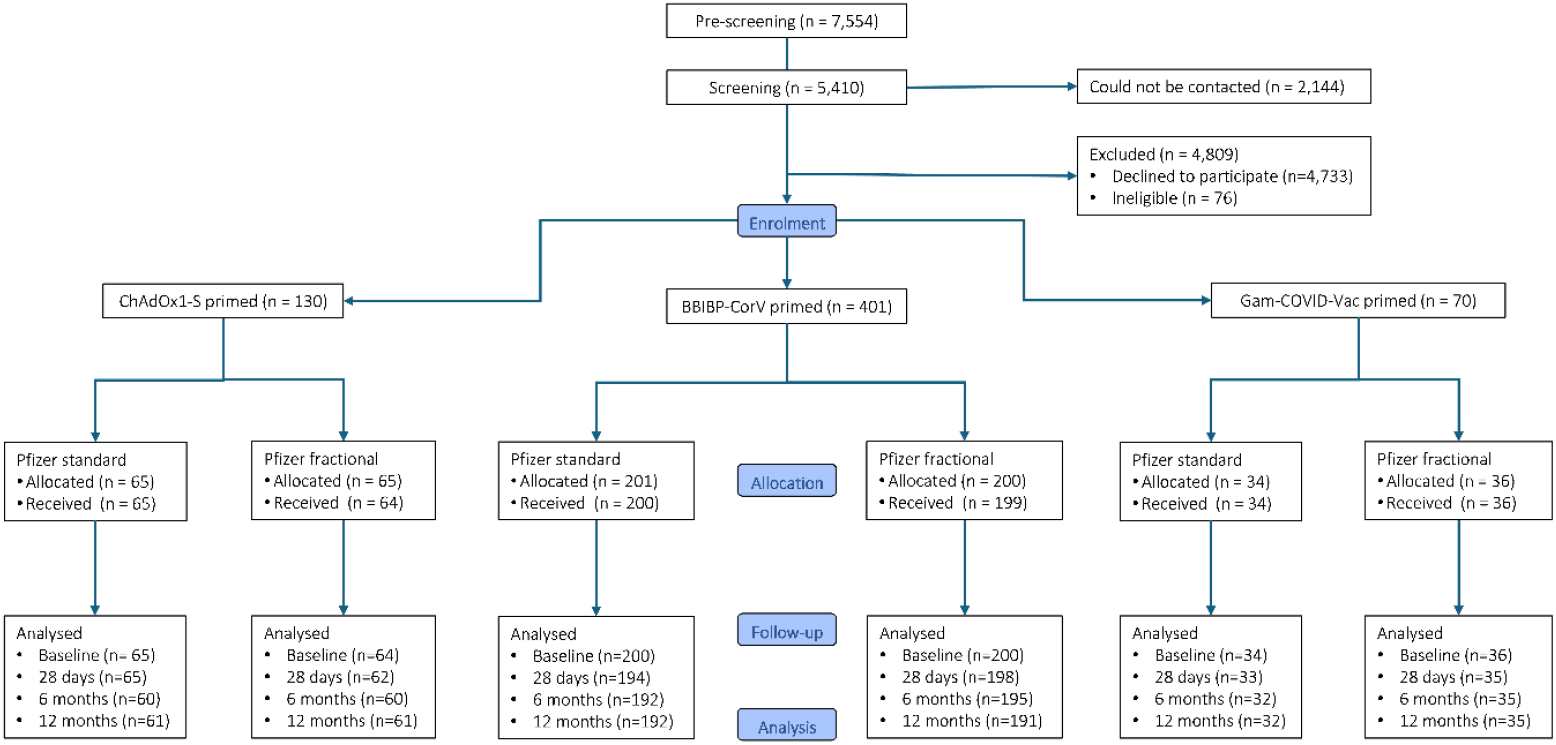
Trial flow-chart stratified by priming vaccine (ChAdOx1-S, BBIBP-CorV, and Gam-COVID-Vac) and study arm (fractional [15μg] or standard [30μg] BNT162b2 booster)

By 12 months, 19 participants had withdrawn. Most withdrawals were due to relocation (n=10) or voluntary withdrawal (n=5). Three withdrawals were due to death: one participant was in the fractional dose arm (primed with BBIBP-CorV), and two participants were in the standard dose arm. One participant was withdrawn as the investigator considered it not in the participant’s best interest to continue (due to a pre-existing medical condition). Two participants were lost to follow-up, one in each study arm, both BBIBP-primed.

### Immunological responses up to 12 months post-booster

#### Anti-spike IgG levels

Missingness for immunological outcomes and covariates for analysis was low (<9% for any variable) (Supplementary Table 2). From 28 days to six months post-booster, anti-spike IgG levels declined in both study arms, then remained stable to 12 months (Supplementary Table 3). IgG levels were similar between study arms at six (GMR 1.03 [95% CI 0.93 – 1.15], *p* = 0.588) and 12 months (1.01 [95% CI 0.90 - 1.14], *p* = 0.834). There were no differences in IgG levels by priming vaccine, except at 12 months among those primed with ChAdOx1-S, where IgG levels in the fractional arm were lower than in the standard dose arm (GMR 0.78 [95% CI 0.63 – 0.96], *p* = 0.017). Results are displayed graphically in Figure 2.

**Figure 2.**
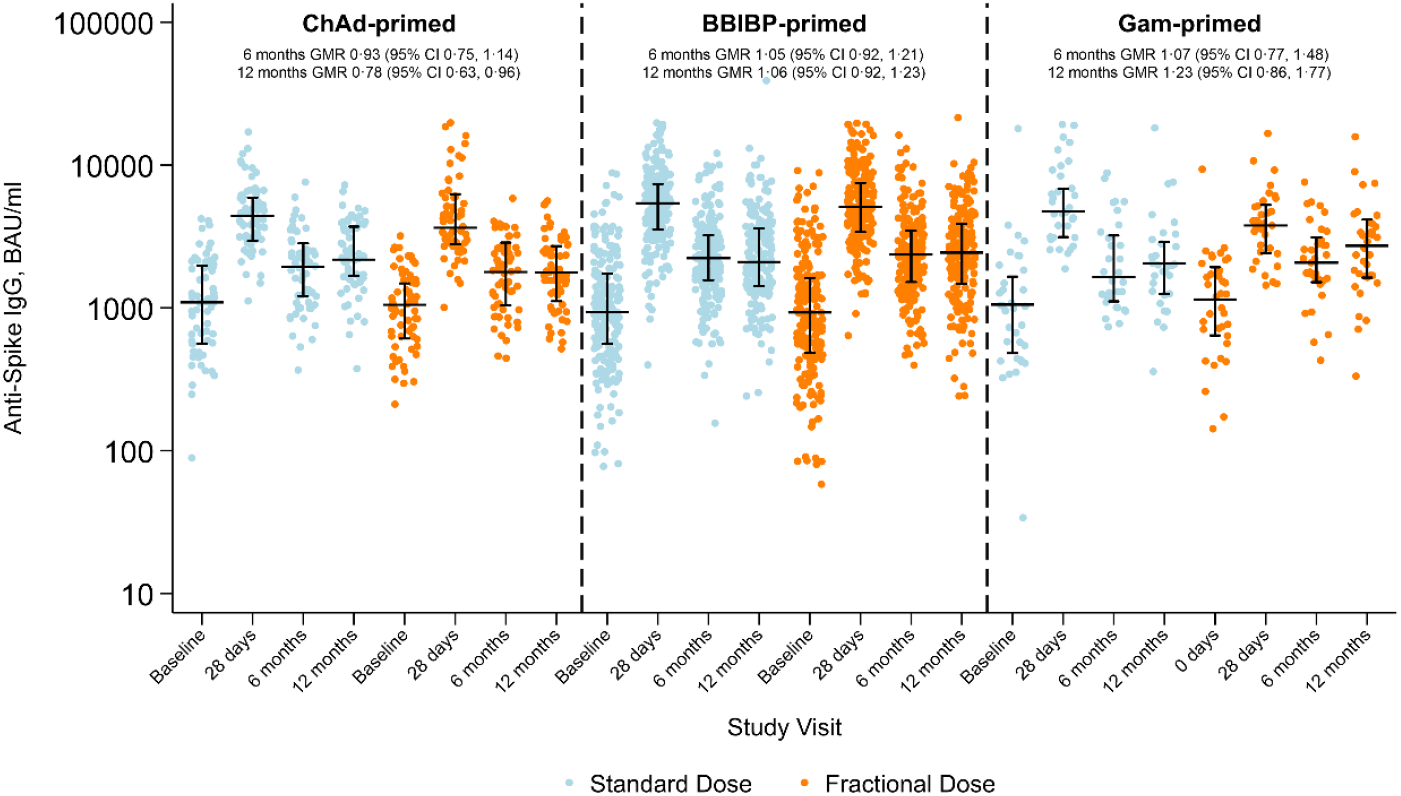
Anti-spike IgG by study visit, priming vaccine, and study arm. Horizontal black lines with vertical bars indicate the median and Interquartile range. GMR: Geometric mean ratio at six and 12months for levels between fractional and standard dose arms, adjusted for age group, priming vaccine, duration between first and second dose, duration between second and third (study) dose, study day of blood draw, and baseline anti-spike IgG

All changes in individual IgG levels between each study visit, study arm, and priming strata are shown in Figure 3. Those with fold changes ≥ 1.2 are highlighted in orange. Many participants also showed decreases in antibody levels after the initial 28-day peak (reflecting the natural waning of immune responses post-vaccination). These results highlight robust early immune responses post-booster, with later increases potentially influenced by SARS-CoV-2 exposure. Additionally, the kinetics of both fractional and standard doses may be similar over time.

**Figure 3.**
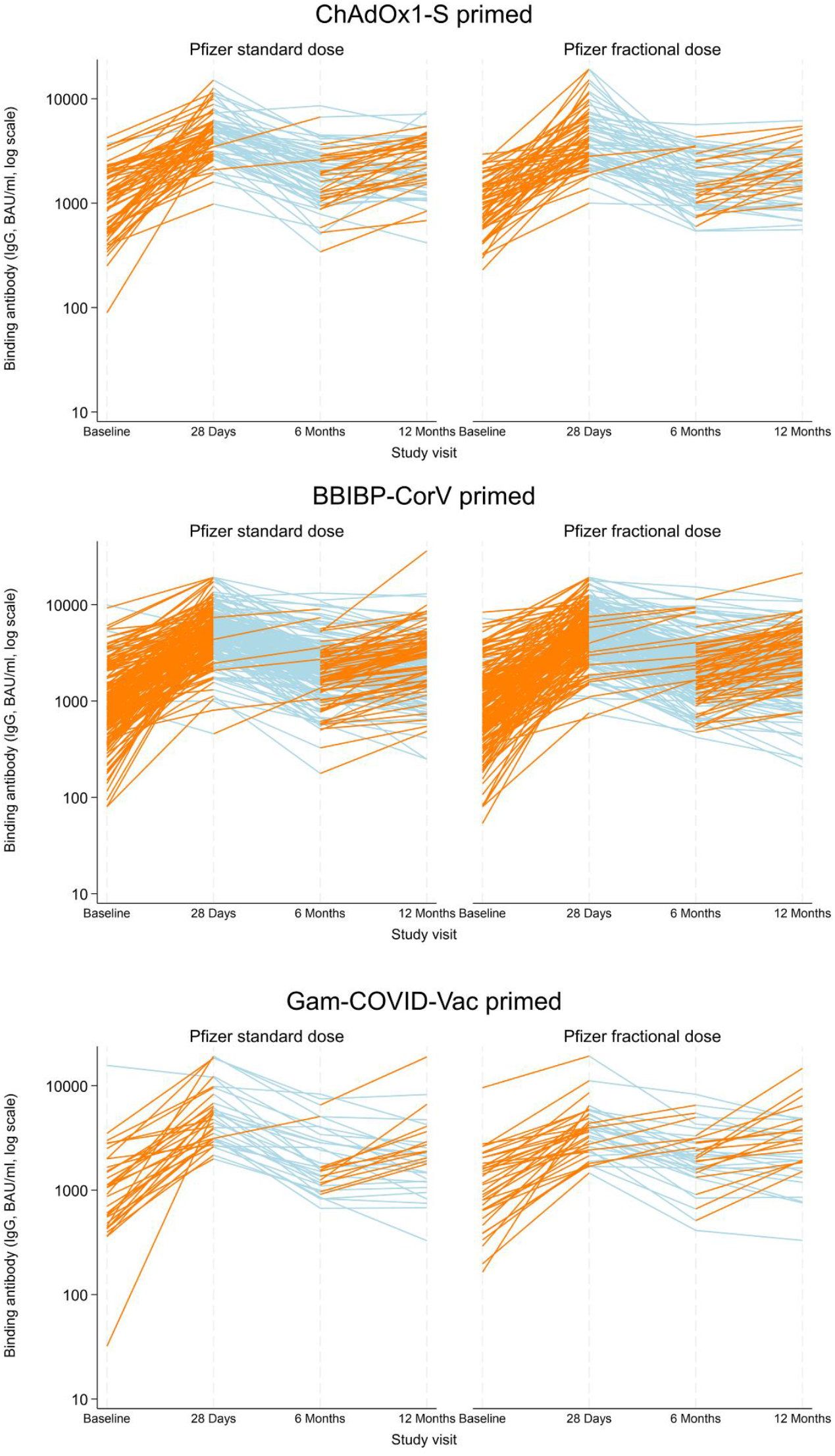
Change in individual binding antibody titres (increase in fold change of ≥ 1.2 in orange, decrease or fold change < 1.2 in light blue) between baseline and 28 days, 28 days and six months, and six and 12 months post-booster, by priming vaccine and study arm

#### Documented and undocumented intercurrent SARS-CoV-2 infections

Table 1 summarises fold changes ≥ 1.2 in anti-spike IgG levels without a documented SARS-CoV-2 infection and documented SARS-CoV-2 infections across study visits, stratified by study arm and priming strata. Documented SARS-CoV-2 infections, shown visually in Supplementary Figure 1, were low throughout the study period, with infection rates of 4.2% (25/601) to 12 months, and with minimal differences between study arms. Undocumented SARS-CoV-2 infections (indicated by a fold change of ≥ 1.2 in anti-spike IgG levels without a documented infection) were higher in the fractional than standard dose arm (20/280 (7.1%) vs 8/271 (3.0%); *p* = 0.025) between Day 28 and six months. Within this interval, participants primed with Gam-COVID-Vac showed higher proportions of fold changes ≥1.2 in the fractional-dose arm compared to the standard-dose arm (6/33 (18.2%) vs. 1/31 (3.2%); *p* = 0.055). Between six and 12 months, a higher proportion of participants with a fold change ≥1.2 were observed in the standard-dose arm among those primed with ChAdOx1-S (27/57 (47.4%) vs. 17/57 (29.8%); *p* = 0.054). These differences may reflect random variation.

**Table 1.** Fold change of ≥ 1.2 in anti-spike levels without documented SARS-CoV-2 infection and documented SARS-CoV-2 infections, per visit by priming vaccine and study arm

| Priming Strata | Fold change $\geq 1.2$ in anti-spike IgG levels without documented SARS-CoV-2 infection n/N<sup>a</sup> (%) | | | Documented SARS-CoV-2 infections, n/N<sup>b</sup> (%) | | |
| --- | --- | --- | --- | --- | --- | --- |
|  | Standard dose | Fractional dose | P-value | Standard dose | Fractional dose | P-value |
| 0 - 28 days<sup>c</sup> |  |  |  |  |  |  |
| All | 282/291 (96.9) | 285/294 (96.9) | 0.982 | 1/292 (0.3) | 1/295 (0.3) | 0.994 |
| ChAdOx1-S | 63/64 (98.4) | 61/62 (98.4) | 0.982 | 1/65 (1.5) | 0/62 (0.0) | 0.327 |
| BBIBP-CorV | 187/194 (96.4) | 189/197 (95.9) | 0.816 | 0/194 (0.0) | 1/198 (0.5) | 0.322 |
| Gam-COVID-Vac | 32/31 (97.0) | 35/35 (100.0) | 0.229 | 0/33 (0.0) | 0/35 (0.0) | - |
| 28 days – 6 months |  |  |  |  |  |  |
| All | 8/271 (3.0) | 20/280 (7.1) | 0.025 | 8/279 (2.9) | 8/290 (2.8) | 0.937 |
| ChAdOx1-S | 2/56 (3.6) | 2/58 (3.5) | 0.972 | 4/60 (6.7) | 2/60 (3.3) | 0.402 |
| BBIBP-CorV | 5/184 (2.7) | 12/189 (6.4) | 0.093 | 4/188 (2.1) | 5/196 (2.6) | 0.784 |
| Gam-COVID-Vac | 1/31 (3.2) | 6/33 (18.2) | 0.055 | 0/31 (0.0) | 1/34 (2.9) | 0.336 |
| Six - 12 months |  |  |  |  |  |  |
| All | 107/275 (38.9) | 92/279 (33.0) | 0.146 | 4/279 (1.4) | 3/283 (1.1) | 0.690 |
| ChAdOx1-S | 27/57 (47.4) | 17/57 (29.8) | 0.054 | 2/59 (3.4) | 1/58 (1.7) | 0.569 |
| BBIBP-CorV | 69/186 (37.1) | 59/187 (31.6) | 0.259 | 2/188 (1.1) | 2/190 (1.1) | 0.992 |
| Gam-COVID-Vac | 11/32 (34.4) | 16/35 (45.7) | 0.345 | 0/32 (0.0) | 0/35 (0.0) | - |

IgG – Immunoglobulin G; Footnotes: <sup>a</sup> Documented SARS-CoV-2 infection excluded from numerator and denominator; <sup>b</sup> Number who attended both Day 0 and Day 28 visits, or Day 28 and six-month visits, or both six and 12 month visits; <sup>c</sup> Documented infections were only considered valid if they occurred between 14 and 28 days, and IgG levels were expected to rise at 0–28 days due to the timing of the measurements shortly after booster administration.

#### Wuhan-Hu-1 SARS-CoV-2 sVNT inhibition

From Day 28 to six months post-booster, the inhibition percentages for Wuhan-Hu-1 SARS-CoV-2 sVNT increased overall from 81 (IQR 78-84) to 89 (IQR 88 - 91) in both study arms (Supplementary Table 4). At twelve months, the inhibition percentages remained stable, with a median of 89% (IQR 88 – 90) for the standard and 89% (IQR 87 – 90) for the fractional study arm, suggesting continued preservation of neutralising activity. The consistency at six and 12 months was observed across all priming strata. Overall, these results indicate that the standard and fractional dosing schedules maintain similar neutralising efficacy against the Wuhan-Hu-1 SARS-CoV-2 strain up to 12 months post-vaccination, regardless of the initial vaccine used for priming (Figure 4).

**Figure 4.**
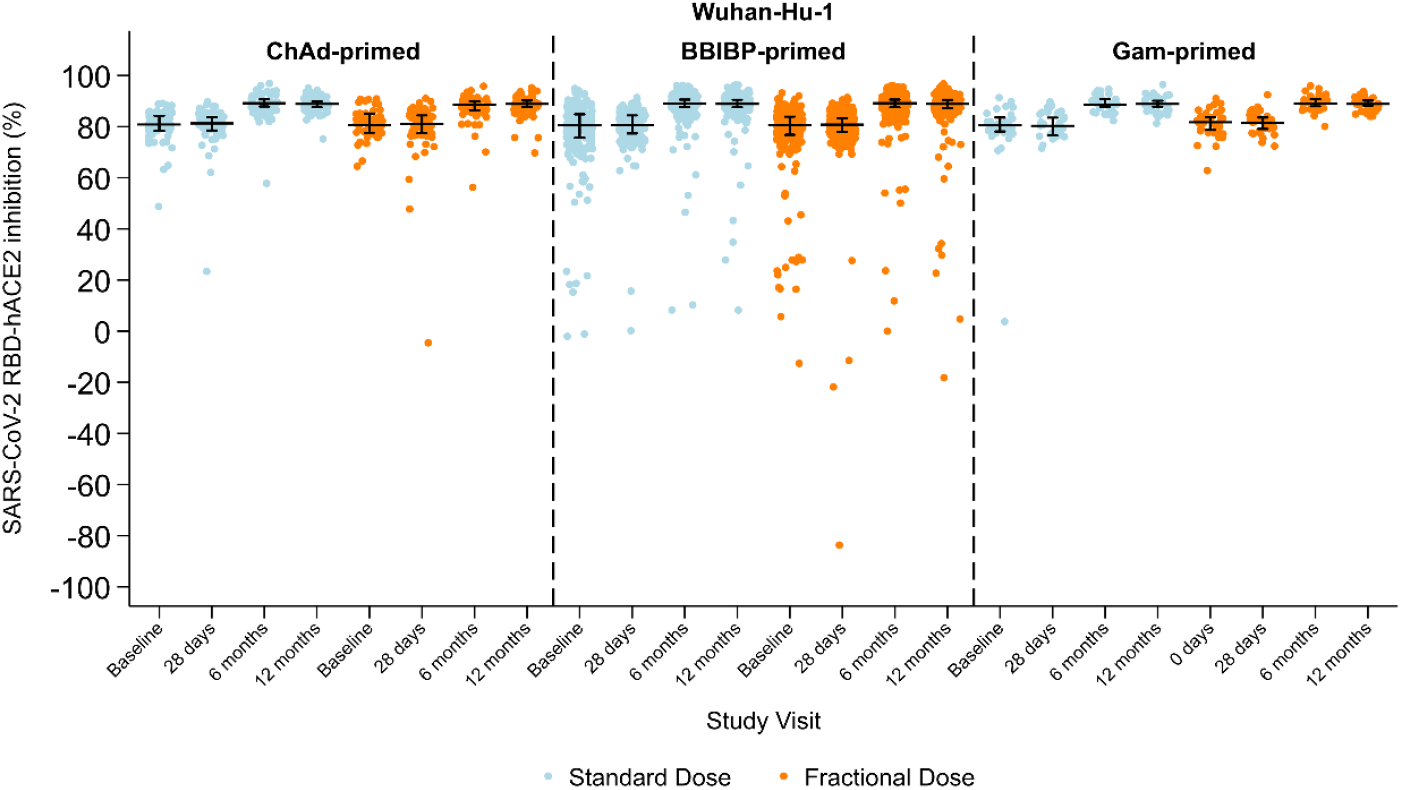
SARS-CoV-2 sVNT inhibition (%) against Wuhan-Hu-1 by study visit, priming vaccine, and study arm. Horizontal black lines with vertical bars indicate the median and interquartile range.

#### Omicron BA.1 SARS-CoV-2 sVNT inhibition

The neutralising response against the Omicron BA.1 variant, measured by sVNT inhibition, shows minimal fluctuations between fractional and standard doses at six and 12 months post-vaccination (Supplementary Table 5 and Figure 5). The median sVNT inhibition percentage against the SARS-CoV-2 Omicron BA.1 showed little change between Day 28 and six months, with the overall median inhibition percentage at six months being 77% (IQR 48 – 87) for the fractional dose and similar at 74% (IQR 46 – 87) for the standard dose. At 12 months, the overall median inhibition percentages were similar for the fractional dose 79% (40 – 87) and standard dose 76% (IQR 44 – 87).

**Figure 5.**
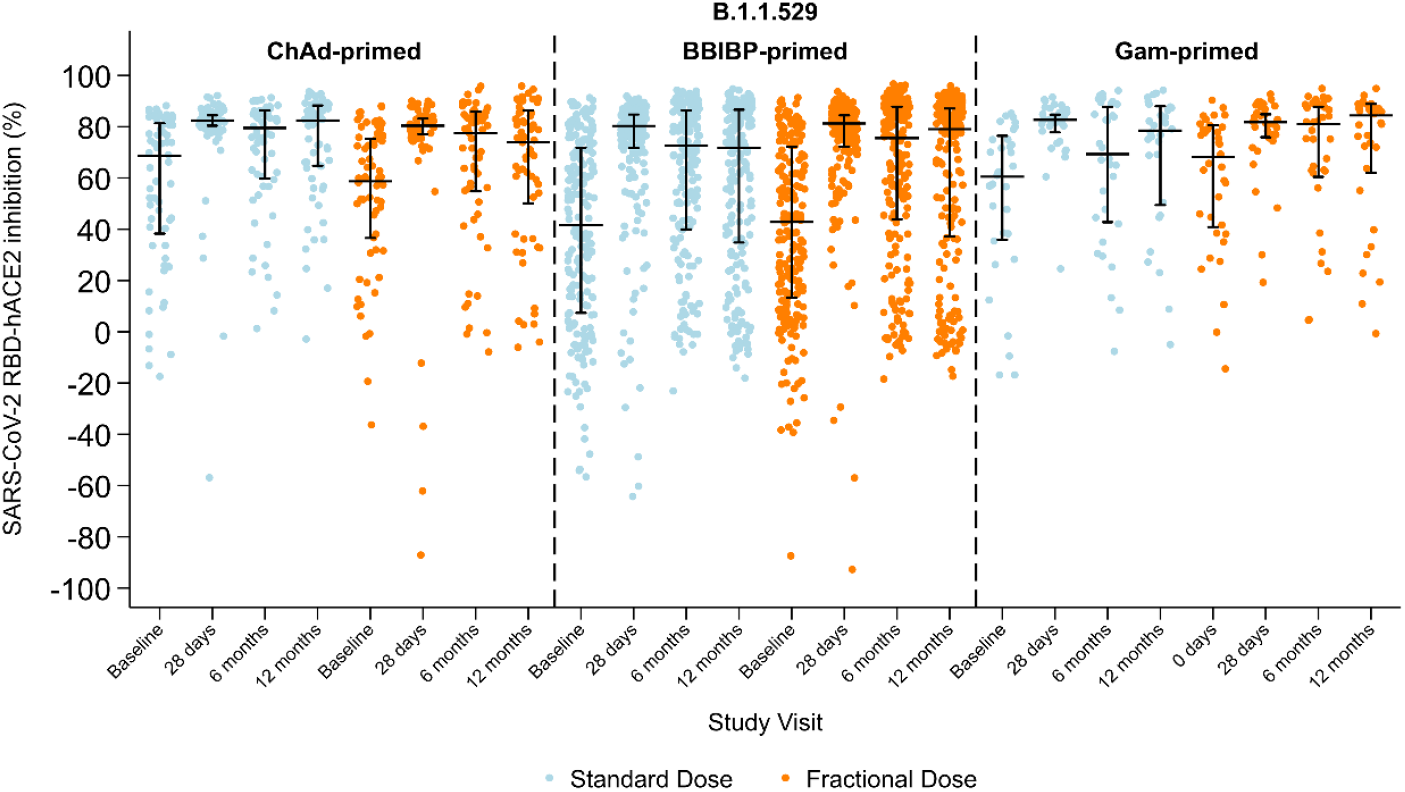
SARS-CoV-2 sVNT inhibition (%) against Omicron BA.1 by study visit, priming vaccine, and study arm. Horizontal black lines with vertical bars indicate the median and interquartile range

By priming strata, the inhibition percentages were similar at six and 12 months. At six months, median inhibition percentages ranged from 76% (IQR 44–88) for BBIBP-CorV to 81% (IQR 60–88) for Gam-COVID-Vac in the fractional dose group and from 73% (IQR 40–86) for BBIBP-CorV to 80% (IQR 60–86) for ChAdOx1-S in the standard dose group. By 12 months, the median inhibition percentages remained similar, with values ranging from 74% (50–86) for ChAdOx1-S to 84% (IQR 62–89) for Gam-COVID-Vac in the fractional dose group, and from 72% (IQR 35–87) for BBIBP-CorV to 82% (IQR 65–88) for ChAdOx1-S in the standard dose group.

### Adverse and serious adverse events

From baseline to the 12 month visit, 76 AEs and 41 SAEs were recorded (Supplementary Tables 6 and 7). The 76 AEs were distributed evenly across study arms, with 38 (50.0%) occurring in each of the fractional and standard groups. Most AEs (48/76, 63.2%) were mild and unrelated to the study vaccine. A smaller proportion (27/76, 35.5%) were classified as possibly related to the vaccine, including irregular menstruation, furuncles, chest pain, and hypertension, which occurred 3 to 83 days post-booster. Most AEs (68/76, 89.5%) resolved completely, while 8/76 (10.5%) resolved with sequelae. Two cases of upper respiratory tract infection were reported and deemed unrelated to the study vaccine.

The SAEs included both acute and chronic medical conditions, with onset times ranging from 1 to 385 days post-vaccination. Most SAEs were moderate (15/41, 36.6%) or severe (21/41, 51.2%), and two (4.9%) were classified as life-threatening. There were three (7.3%) fatal outcomes, attributed to suicide, decompensated diabetes, and gastric cancer. Among severe SAEs, 12 resolved with sequelae, while the remaining nine resolved without long-term effects. Of the 41 SAEs, 21 (51.2%) occurred in the fractional dose group and 20 (48.8%) in the standard dose group. Importantly, no severe vaccine-related AEs or SAEs were reported.

## Discussion

This study assessed the longitudinal immunogenicity and safety of fractional and standard BNT162b2 booster doses over 12 months in Mongolian adults primed with ChAdOx1-S, BBIBP-CorV, or Gam-COVID-Vac. It provides the first long-term follow-up data on booster strategies for populations primed with inactivated (BBIBP-CorV) or viral vector vaccines (ChAdOx1-S and Gam-COVID-Vac) in the region, which is particularly relevant for low- and middle-income countries, where these vaccines are used widely. Our earlier findings demonstrated that fractional BNT162b2 boosters were non-inferior to standard doses for anti-spike IgG seroconversion in ChAdOx1-S and BBIBP-CorV primed strata, though responses were lower in the Gam-COV-Vac stratum at one-month post-vaccination.[2] Over 12 months, both fractional and standard BNT162b2 booster doses maintained similar anti-spike IgG levels, except for the ChAdOx1-S stratum, where the fractional dose showed lower levels. Neutralising antibody responses against the Wuhan-Hu-1 strain remained stable across study arms and by priming strata, while responses to the Omicron BA.1 declined at six and 12 months but remained above baseline, indicating lasting cross-protection to this variant. SARS-CoV-2 infections were similar between groups, and no severe vaccine-related adverse events were observed, supporting the safety and potential of fractional dose use in resource-constrained settings.

Anti-spike IgG levels were sustained over 12 months post-booster, with some variability by priming vaccine. The variability may reflect slower decay rates observed in humoral responses following viral vector vaccine boosters compared with mRNA boosters, particularly in heterologous schedules.[10] Our findings align with a 2022 systematic review of 28 studies (5870 subjects), which found that booster vaccination significantly increased anti-spike IgG levels, with heterologous combinations—especially inactivated-RNA combinations —showing superior responses.[11] mRNA boosters (BNT162b2 and mRNA-1273) demonstrated durability, maintaining 1.5– 3.4 times higher IgG levels at six months.[11] An observational study in the United States of 55 adults (ages 18– 50) showed a 21-fold increase in IgG after the third dose of mRNA vaccine (BNT162b2 or mRNA1273), with a GMT of 9.18 × 10^5^ AU/mL at one month, declining 3-fold to 3 × 10^5^ AU/mL at six months, with no significant differences between mRNA vaccine types.[12] In this study, in Mongolian adults, following a five-fold increase in IgG levels post-booster for both study arms, IgG levels declined approximately 2.5-fold from 28 days to six months and then remained stable through to 12 months. Few studies, however, have explored fractional dosing beyond six months. Our results align with the COV-BOOST trial in the UK, which found similar IgG responses between half and full doses of BNT162b2 at one, three, and eight months, highlighting the importance of tailored booster strategies that consider variations in initial vaccine platforms to optimise long-term immune responses.[4, 10]

Neutralising antibody responses in our Mongolian cohort, as measured by sVNT inhibition, were stable over time for Wuhan-Hu-1 strain while demonstrating cross-reactivity against the Omicron BA.1 variant, further highlighting the long-term protective potential of both dosing regimens. While binding antibody levels to Omicron BA.1 were not measured in our study, evidence suggests these responses are typically lower than those to ancestral strains, despite the presence of functional neutralising activity observed with sVNT.[2, 13] Future research should explore this dynamic further to optimise booster strategies against emerging variants.

Our findings align with global evidence supporting tailored strategies, relevant to Mongolia and similar settings, where a range of non-mRNA vaccines have been used as primary doses. A pooled analysis of 14 studies found that variant-modified boosters elicited a 1.6-fold higher neutralising antibody titre against Omicron subvariants compared to ancestral-based vaccines, particularly when the booster closely matched the circulating variant, highlighting the value of tailored strategies.[14] The PICOBOO trial demonstrated that mRNA boosters (BNT162b2 and mRNA-1273) produced higher initial binding and neutralising titres than NVX-CoV2373 (a protein subunit vaccine), though NVX-CoV2373 maintained more durable responses over three months, suggesting its potential for long-term protection, particularly in contexts where durability is a key consideration.[15] Similarly, the COVAIL study found robust responses from bivalent mRNA boosters targeting Omicron BA.1 or BA.4/BA.5, though efficacy against newer variants like XBB.1 and BQ.1.1 was reduced.[16] These results underscore the challenges of antigenic mismatch and the need for adaptable strategies. While our study demonstrates that fractional doses may offer similar protection to standard doses against earlier SARS-CoV-2 variants, further research is needed to determine whether this holds for newer variants, particularly in terms of long-term effectiveness and cross-protection.

The incidence of documented and undocumented SARS-CoV-2 breakthrough infections during follow-up periods provides critical insights into the interplay between booster dosing regimens and real-world protection against evolving variants. The reported incidence of SARS-CoV-2 infections among study participants was low, while immune response patterns suggest a substantial number of additional undocumented cases. However, increases in IgG titres were observed in both arms, albeit more frequently in the fractional dose arm, indicating a high incidence of intercurrent SARS-CoV-2 infection with low case ascertainment. Our findings between six and 12 months, where a subset of participants experienced increased IgG titres without documented infections, highlight the limitations of ongoing COVID-19 surveillance to accurately monitor SARS-CoV-2 infections, particularly in the context of low community-level testing. These data are consistent with many studies that highlight discrepancies between self-reporting and serological surveillance, suggesting that self-reported cases often underestimate true infection rates.[12, 14-17] Despite regular follow-up and requests for participants to report positive test results and symptoms, the reliance on self-reported testing and the absence of anti-nucleocapsid protein testing led to under-ascertainment of true infection rates in our study.[18] This limitation underscores the challenge of monitoring breakthrough infections in the context of waning community-level testing and surveillance infrastructure, particularly as attention shifts from acute case detection to long-term disease management.

However, several limitations should be considered. Our SARS-CoV-2 spike-specific IgG antibody results were only evaluated against Wuhan, and our neutralising results were only measured to Wuhan and Omicron BA.1. While our results have provided valuable long-term insights into the immune response of fractional dosing, there would be added value of evaluating the cross-protection to newer circulating SARS-CoV-2 variants. The reliance on Spike IgG titre changes to infer undocumented infections, without anti-nucleocapsid testing, is a key limitation.[18] This approach may underestimate true infection rates, however since we have a Sinopharm-primed group that used the inactivated virus in the vaccine, anti-nucleocapsid testing would not be reliable.

These evaluations are ongoing and will provide further insight into the effectiveness of these booster strategies against emerging variants. Finally, the study was conducted in a single country (Mongolia), and while similar studies may provide complementary insights, results from our study may not be generalisable to other populations with different demographic or epidemiological profiles.[19]

This study’s strengths include its randomised, double-blind design and extended follow-up period of 12 months, allowing for comprehensive assessment of long-term immune responses. Including participants primed with different vaccines (ChAdOx1-S, BBIBP-CorV, and Gam-COVID-Vac) provides a broad perspective on the effectiveness of fractional doses across various vaccination backgrounds. This diversity is particularly relevant for many LMICs that have used these vaccines extensively, making the findings applicable to diverse global populations. Additionally, the study achieved a high retention rate (94%) over twelve months, further strengthening the robustness of the findings. This study is ongoing, collecting data up to 24 months, and will be important for understanding long-term immunity following fractional dosing, including cellular mediated immunity against emerging variants. This is particularly relevant for many LMICs that have only administered three doses of COVID-19 vaccines and have limited access to next generation monovalent variant vaccines.

## Conclusion

This study demonstrates that fractional doses of the BNT162b2 vaccine can elicit sustained and robust immune responses comparable to standard doses up to 12 months post-booster, supporting their potential use in optimising booster vaccination strategies. Our findings provide critical insights into the long-term performance of fractional dosing and contribute to the broader understanding of effective booster strategies in populations primed with diverse COVID-19 vaccines. Further research is needed to explore the broader applicability of these results in other settings and to evaluate the impact of undocumented SARS-CoV-2 infections on the breadth and duration of protective immunity.

## Data Availability

All data produced in the present study are available upon reasonable request to the authors

https://clinicaltrials.gov/study/NCT05265065

https://pubmed.ncbi.nlm.nih.gov/38357398/

## Data Availability

All data produced in the present study are available upon reasonable request to the authors

https://clinicaltrials.gov/study/NCT05265065

https://pubmed.ncbi.nlm.nih.gov/38357398/

## Acknowledgements

This study was made possible by the generosity of the study participants – we thank them for their time and cooperation in the study procedures. We would like to acknowledge the invaluable contributions of: Dr Chinburen Jigjidsuren (Member of Parliament); Dr Ganzorig Dorjdagva (Ministry of Health); Dr Urangoo Khurlee, Dr Gentsenpilmaa Batbold, Dr Gantuya Damdinsuren, and Dr Altanshahai Boldbaatar (First Central Hospital of Mongolia); Dr Tserendagva Dalkh (Mongolian National University of Medical Sciences); Dr Davaalkham Jagdagsuren, Dr Naranzul Tsedenbal, Tserendulam Bazarkhuu, and Ariunbileg Gankhuyag (National Centre for Communicable Diseases); Dr Budkhand Ichinkhorloo, and Dr Erdenebayar Tsedevsuren (Onoshmed Laboratory); Dr Baldandugar Zeeren and Dr Davaajav Tsend-Ayush (Bayangol District Health Department); Bat-Ireedui Purevbaatar and Dr Gantuya Gansukh (Sukhbaatar District Health Department); Dr Davaajargal Oyunsuren (Songinokhairkhan District Health Department); Dr Gandiimaa Riimaadai, Dr Tserendulam Dorjsuren and Dr Ariunaa Jadambaa (Arkhangai Province Health Department); and Dr Erkegul Sandalhan (Railway Central Hospital). This clinical trial is funded by CEPI, an innovative global partnership working to accelerate the development of vaccines against epidemic and pandemic threats so they can be accessible to all people in need. Grant management support is provided by PATH, an international non-profit global health organization. The Government of Mongolia provided the study vaccines. Kerryn Moore is supported by an Australian National Health and Medical Research Council (NHMRC) Early Career Fellowship (Grant Number APP1160936); the content of this publication is solely the responsibility of the authors and does not necessarily represent the official views of the NHMRC. Authors affiliated with MCRI were supported by the Victorian Government’s Operational Infrastructure Support Program.

## Data sharing statement

Anonymous participant data that underlie the results reported in this Article will be available on completion of the clinical trial. Data requests should be sent to the corresponding author. The requester must provide a scientifically sound proposal and data transfer agreement for the sponsors’ and collaborators’ approval. On approval, data will be transferred through a secure online platform.

