## Supplementary Materials for "Twelve-month follow-up of immunogenicity and safety of fractional and standard booster doses of the Pfizer-BioNTech COVID-19 vaccine in adults primed with ChAdOx1, BBIBP-CorV, or GAM-CoV-Vac: a randomised controlled trial"

### Participant recruitment and follow-up

Supplementary Table 1 presents the baseline characteristics of 598 participants who received the intervention, stratified by vaccine allocation (standard vs. fractional dose) and initial priming vaccine (ChAdOx1-S, BBIBP-CorV, Gam-COVID-Vac), as described previously.[1] Here, we summarise self-reported infection with SARS-CoV-2 before study commencement. Overall, 49.2% of participants reported prior SARS-CoV-2 infection. ChAdOx1-S primed participants had infection rates of 60.0% (standard dose) and 64.1% (fractional dose); BBIBP-CorV primed participants had rates of 45.5% (standard) and 37.2% (fractional); Gam-COVID-Vac primed participants reported the highest prior infection rates, with 58.8% (standard) and 80.6% (fractional).

*Supplementary Table 1 Baseline characteristics by study vaccine allocation and priming strata*

| | **All Priming Strata** | | | **Primed with ChAdOx1-S** | | **Primed with BBIBP-CorV** | | **Primed with Gam-COVID-Vac** | |
|---|---|---|---|---|---|---|---|---|---|
| | **Total** | **Standard** | **Fractional** | **Standard** | **Fractional** | **Standard** | **Fractional** | **Standard** | **Fractional** |
| | **n = 598** | **n = 299** | **n = 299** | **n = 65** | **n = 64** | **n = 200** | **n = 199** | **n = 34** | **n = 36** |
| Age, years, median (IQR) | 44 (32-55) | 44 (32-55) | 44 (33-55) | 34 (32-46) | 40 (34-50) | 48 (31-58) | 48 (32-57) | 43 (32-53) | 41 (35-50) |
| <50 years | 360 (60.2%) | 181 (60.5%) | 179 (59.9%) | 54 (83.1%) | 50 (78.1%) | 103 (51.5%) | 103 (51.8%) | 24 (70.6%) | 26 (72.2%) |
| ≥50 years | 238 (39.8%) | 118 (39.5%) | 120 (40.1%) | 11 (16.9%) | 14 (21.9%) | 97 (48.5%) | 96 (48.2%) | 10 (29.4%) | 10 (27.8%) |
| Male sex | 273 (45.7%) | 132 (44.1%) | 141 (47.2%) | 32 (49.2%) | 33 (51.6%) | 85 (42.5%) | 86 (43.2%) | 15 (44.1%) | 22 (61.1%) |
| Female sex | 325 (54.3%) | 167 (55.9%) | 158 (52.8%) | 33 (50.8%) | 31 (48.4%) | 115 (57.5%) | 113 (56.8%) | 19 (55.9%) | 14 (38.9%) |
| BMI, kg/m$^{2}$, | 25.2 (22.6-28.7) | 25.2 (22.7-28.9) | 25.1 (22.5-28.7) | 26.3 (23.7-30) | 25.4 (23.4-28.4) | 24.6 (22.0-28.7) | 24.8 (22.3-27.9) | 25.3 (24.2-28.7) | 25.8 (23.1-29.8) |
| Days between 1$^{st}$ and 2$^{nd}$ doses, median (IQR) | 31 (24-45) | 30 (24-44) | 31 (24-46) | 43 (41-50) | 44 (41-51) | 28 (23-30) | 28 (23-33) | 58 (44-64) | 61 (58-64) |
| Days between 2$^{nd}$ dose and study (3$^{rd}$) dose | 428 (397-454) | 432 (391-454) | 425 (400-454) | 494 (416-517) | 505 (410-519) | 422 (386-450) | 418 (397-450) | 419 (367-448) | 430 (388-450) |
| Reaction following 1st or 2nd dose | 93 (15.6%) | 45 (15.1%) | 48 (16.1%) | 21 (32.3%) | 18 (28.1%) | 17 (8.5%) | 24 (12.1%) | 7 (20.6%) | 6 (16.7%) |
| Pain or fever medication taken for reaction | 21 (22.6%) | 9 (20.0%) | 12 (25.0%) | 7 (33.3%) | 8 (44.4%) | 1 (5.9%) | 4 (16.7%) | 1 (14.3%) | 0 (0.0%) |
| Medical advice sought for reaction | 3 (3.2%) | 1 (2.2%) | 2 (4.2%) | 1 (4.8%) | 1 (5.6%) | 0 (0.0%) | 1 (4.2%) | 0 (0.0%) | 0 (0.0%) |
| Symptoms of reaction resolved | 53 (57.0%) | 28 (62.2%) | 25 (52.1%) | 11 (52.4%) | 5 (27.8%) | 10 (58.8%) | 15 (62.5%) | 7 (100%) | 5 (83.3%) |
| Self-reported prior SARS-CoV-2 infection before study commencement | 294 (49.2%) | 150 (50.2%) | 144 (48.2%) | 39 (60.0%) | 41 (64.1%) | 91 (45.5%) | 74 (37.2%) | 20 (58.8%) | 29 (80.6%) |
| Comorbidities | | | | | | | | | |
| Obesity (BMI ≥30 kg/m$^{2}$) | 115 (19.2%) | 55 (18.4%) | 60 (20.1%) | 13 (20.0%) | 12 (18.8%) | 37 (18.5%) | 39 (19.6%) | 5 (14.7%) | 9 (25.0%) |
| Diabetes mellitus | 25 (4.2%) | 17 (5.7%) | 8 (2.7%) | 2 (3.1%) | 1 (1.6%) | 10 (5.0%) | 5 (2.5%) | 5 (14.7%) | 2 (5.6%) |
| Cardiovascular disease | 56 (9.4%) | 26 (8.7%) | 30 (10.0%) | 2 (3.1%) | 5 (7.8%) | 20 (10.0%) | 23 (11.6%) | 4 (11.8%) | 2 (5.6%) |
| Hypertension | 166 (27.8%) | 80 (26.8%) | 86 (28.8%) | 14 (21.5%) | 14 (21.9%) | 59 (29.5%) | 60 (30.2%) | 7 (20.6%) | 12 (33.3%) |
| Cancer | 3 (0.5%) | 0 (0.0%) | 3 (1.0%) | 0 (0.0%) | 0 (0.0%) | 0 (0.0%) | 1 (0.5%) | 0 (0.0%) | 2 (5.6%) |

| | | | | | | | | | |
|---|---|---|---|---|---|---|---|---|---|
| Chronic obstructive pulmonary disease | 8 (1.3%) | 4 (1.3%) | 4 (1.3%) | 0 (0.0%) | 0 (0.0%) | 2 (1.0%) | 3 (1.5%) | 2 (5.9%) | 1 (2.8%) |
| Chronic kidney disease | 49 (8.2%) | 25 (8.4%) | 24 (8.0%) | 3 (4.6%) | 4 (6.2%) | 18 (9.0%) | 17 (8.5%) | 4 (11.8%) | 3 (8.3%) |
| Chronic liver disease | 19 (3.2%) | 9 (3.0%) | 10 (3.3%) | 1 (1.5%) | 3 (4.7%) | 6 (3.0%) | 4 (2.0%) | 2 (5.9%) | 3 (8.3%) |
| History of anaphylaxis (or carry EpiPen) | 12 (2.0%) | 6 (2.0%) | 6 (2.0%) | 3 (4.6%) | 2 (3.1%) | 3 (1.5%) | 3 (1.5%) | 0 (0.0%) | 1 (2.8%) |
| Neurological disease (including stroke) | 6 (1.0%) | 3 (1.0%) | 3 (1.0%) | 1 (1.5%) | 1 (1.6%) | 2 (1.0%) | 2 (1.0%) | 0 (0.0%) | 0 (0.0%) |
| On anticoagulant therapy | 33 (5.5%) | 17 (5.7%) | 16 (5.4%) | 4 (6.2%) | 2 (3.1%) | 9 (4.5%) | 11 (5.5%) | 4 (11.8%) | 3 (8.3%) |
| Immunocompromised | 0 (0.0%) | 0 (0.0%) | 0 (0.0%) | 0 (0.0%) | 0 (0.0%) | 0 (0.0%) | 0 (0.0%) | 0 (0.0%) | 0 (0.0%) |
| Mastocytosis causing recurrent anaphylaxis | 1 (0.2%) | 0 (0.0%) | 1 (0.3%) | 0 (0.0%) | 0 (0.0%) | 0 (0.0%) | 1 (0.5%) | 0 (0.0%) | 0 (0.0%) |
| Cigarette user | 125 (20.9%) | 66 (22.1%) | 59 (19.7%) | 16 (24.6%) | 17 (26.6%) | 37 (18.5%) | 34 (17.1%) | 13 (38.2%) | 8 (22.2%) |
| Currently pregnant | 1 (0.2%) | 1 (0.3%) | 0 (0.0%) | 1 (1.5%) | 0 (0.0%) | 0 (0.0%) | 0 (0.0%) | 0 (0.0%) | 0 (0.0%) |

Data are median (IQR) or n (%). No data for reported variables were missing. This table excludes three participants who withdrew before receiving the study vaccine.

#### *Missingness of immunological outcomes and covariates for immunogenicity analyses*

Supplementary Table 2 presents the levels of missing data for visit attendance and covariates by study visit, vaccine allocation, and priming strata. Missingness for anti-Spike IgG levels was low across all time points, with 0.3% (2/601) missing at Day 0, 2.3% (14/601) at Day 28, 4.5% (27/601) at six months, and 4.5% (27/601) at 12 months (Supplementary Table 2). The same rates of missingness were observed for sVNT inhibition percentages. When stratified by study arm and priming vaccine, missingness was comparable, with slight variations across strata that did not suggest systematic differences, suggesting missingness was not substantially influenced by either the dosing strategy or priming vaccine.

Missing data for covariates included in linear regression models built to assess long-term immunogenicity were minimal and evenly distributed across study arms and priming strata (Supplementary Table 2). Age group and priming vaccine data were fully complete (0.0% missingness). Dates of first and second vaccine doses were entirely recorded, while the date of the third (study) dose was missing for 3/601 (0.5%) of participants, representing the three participants who did not receive their allocated study intervention. The study day of blood draw had up to 4.5% missingness at the 12-month visit, with similar rates between standard and fractional dose groups, indicating negligible bias in sample timing analysis. Baseline anti-spike IgG levels (Day 0) were missing for 0.2% of participants (representing one participant who was withdrawn as the investigator considered it not in their best interest to continue). The low level of missingness across covariates suggests that the impact on the immunogenicity analyses was minimal, preserving the validity and robustness of the study findings.

*Supplementary Table 2 Table of missingness for anti-Spike IgG levels, inhibition percentages of Wuhan-Hu-1 and Omicron B.1.1.529 (BA.1) surrogate virus neutralising test and covariates for immunogenicity analysis by study arm and priming strata among all randomised participants*

| | **All Priming Strata** | | | **Primed with ChAdOx1-S** | | **Primed with BBIBP-CorV** | | **Primed with Gam-COVID-Vac** | |
|---|---|---|---|---|---|---|---|---|---|
| **Variable** | **Total (N = 601)** | **Standard (N = 300)** | **Fractional (N= 301)** | **Standard (N = 65)** | **Fractional (N = 65)** | **Standard (N = 201)** | **Fractional (N = 200)** | **Standard (N = 34)** | **Fractional (N = 36)** |
| | n/N (%) | n/N (%) | n/N (%) | n/N (%) | n/N (%) | n/N (%) | n/N (%) | n/N (%) | n/N (%) |
| Anti-Spike IgG levels | | | | | | | | | |
| Day 0 | 2 (0.3) | 1 (0.3) | 1 (0.3) | 0 (0.0) | 1 (1.5) | 1 (0.5) | 0 (0.0) | 0 (0.0) | 0 (0.0) |
| Day 28 | 14 (2.3) | 8 (2.7) | 6 (2.0) | 0 (0.0) | 3 (4.6) | 7 (3.5) | 2 (1.0) | 1 (2.9) | 1 (2.8) |
| Six months | 27 (4.5) | 16 (5.3) | 11 (3.7) | 5 (7.7) | 5 (7.7) | 9 (4.5) | 5 (2.5) | 2 (5.9) | 1 (2.8) |
| 12 months | 27 (4.5) | 14 (4.7) | 13 (4.3) | 4 (6.2) | 4 (6.2) | 9 (4.5) | 8 (4.0) | 1 (3.0) | 1 (2.8) |
| sVNT inhibition (Wuhan-Hu-1 and Omicron BA.1) | | | | | | | | | |
| Day 0 | 2 (0.3) | 1 (0.3) | 1 (0.3) | 0 (0.0) | 1 (1.5) | 1 (0.5) | 0 (0.0) | 0 (0.0) | 0 (0.0) |
| Day 28 | 14 (2.3) | 8 (2.7) | 6 (2.0) | 0 (0.0) | 3 (4.6) | 7 (3.5) | 2 (1.0) | 1 (2.9) | 1 (2.8) |
| Six months | 27 (4.5) | 16 (5.3) | 11 (3.7) | 5 (7.7) | 5 (7.7) | 9 (4.5) | 5 (2.5) | 2 (5.9) | 1 (2.8) |
| 12 months | 32 (5.3) | 17 (5.7) | 15 (5.0) | 4 (6.2) | 4 (6.2) | 11 (5.5) | 9 (4.5) | 2 (6.1) | 2 (5.6) |
| Age-group | 0 (0.0) | 0 (0.0) | 0 (0.0) | 0 (0.0) | 0 (0.0) | 0 (0.0) | 0 (0.0) | 0 (0.0) | 0 (0.0) |
| Priming vaccine | 0 (0.0) | 0 (0.0) | 0 (0.0) | 0 (0.0) | 0 (0.0) | 0 (0.0) | 0 (0.0) | 0 (0.0) | 0 (0.0) |
| Date dose 1 received | 0 (0.0) | 0 (0.0) | 0 (0.0) | 0 (0.0) | 0 (0.0) | 0 (0.0) | 0 (0.0) | 0 (0.0) | 0 (0.0) |
| Date dose 2 received | 0 (0.0) | 0 (0.0) | 0 (0.0) | 0 (0.0) | 0 (0.0) | 0 (0.0) | 0 (0.0) | 0 (0.0) | 0 (0.0) |
| Date dose 3 received[a] | 3 (0.5) | 1 (0.3) | 2 (0.3) | 0 (0.0) | 1 (1.5) | 1 (0.5) | 1 (0.5) | 0 (0.0) | 0 (0.0) |
| Study day of blood draw | | | | | | | | | |
| Day 0 | 1 (0.2) | 1 (0.3) | 0 (0.0) | 0 (0.0) | 0 (0.0) | 1 (0.5) | 0 (0.0) | 0 (0.0) | 0 (0.0) |
| Day 28 | 14 (2.3) | 8 (2.7) | 6 (2.0) | 0 (0.0) | 3 (4.6) | 7 (3.5) | 2 (1.0) | 1 (2.9) | 1 (2.8) |
| Six months | 27 (4.5) | 16 (5.3) | 11 (3.7) | 5 (7.7) | 5 (7.7) | 9 (4.5) | 5 (2.5) | 2 (5.9) | 1 (2.8) |
| 12 months | 27 (4.5) | 14 (4.7) | 13 (4.3) | 4 (6.2) | 4 (6.2) | 9 (4.5) | 8 (4.0) | 1 (3.0) | 1 (2.8) |

[a] Three participants did not receive their allocated investigational product.

### Immunological responses up to 12 months post-vaccination

#### *Anti-spike IgG levels*

Supplementary Table 3 summarises the GMC of anti-Spike IgG antibodies in each study arm and the GMR comparing the fractional and standard dose arms at all timepoints.

*Supplementary Table 3 Geometric mean anti-spike IgG and the geometric mean ratio at baseline, day 28, six- and twelve-month visits by study arm and priming strata.*

| Priming Strata | GMC IgG (95% CI) Standard Dose | GMC IgG (95% CI) Fractional Dose | GMR fractional/standard (95% CI) p- value |
|---|---|---|---|
| **Baseline** | | | |
| All | 969 (876, 1072) [n = 299] | 929 (839, 1029) [n = 300] | 0.94 (0.82, 1.08) p = 0.398 |
| ChAdOx1-S | 1023 (845, 1238) [n = 65] | 958 (820, 1119) [n = 64] | 0.94 (0.74, 1.20) p = 0.620 |
| BBIBP-CorV | 956 (843, 1084) [n = 200] | 899 (785, 1031) [n = 200] | 0.92 (0.77, 1.10) p = 0.370 |
| Gam-COVID-Vac | 953 (667, 1360) [n = 34] | 1052 (788, 1404) [n = 36] | 1.09 (0.72, 1.65) p = 0.685 |
| ***Day 28*** | | | |
| All | 4946 (4614, 5302) [n = 292] | 4619 (4292, 4970) [n = 295] | 0.95 (0.86, 1.04); p = 0.237 |
| ChAdOx1-S | 4394 (3863, 4997) [n = 65] | 4167 (3550, 4892) [n = 62] | 0.94 (0.77, 1.14); p = 0.530 |
| BBIBP-CorV | 5109 (4678, 5580) [n = 194] | 4970 (4541, 5438) [n = 198] | 0.99 (0.88, 1.11); p = 0.853 |
| Gam-COVID-Vac | 5160 (4128, 6450) [n = 33] | 3662 (3016, 4447) [n = 35] | 0.71 (0.55, 0.91); p = 0.009 |
| ***Six months*** | | | |
| All | 2085 (1926, 2256) [n = 284] | 2123 (1964, 2296) [n = 290] | 1.03 (0.93, 1.15); p = 0.588 |
| ChAdOx1-S | 1832 (1556, 2157) [n = 60] | 1706 (1466, 1985) [n = 60] | 0.93 (0.75, 1.14); p = 0.471 |
| BBIBP-CorV | 2195 (1991, 2419) [n = 192] | 2286 (2074, 2521) [n = 195] | 1.05 (0.92, 1.21); p = 0.441 |
| Gam-COVID-Vac | 1950 (1511, 2517) [n = 32] | 2044 (1628, 2569) [n = 35] | 1.07 (0.77, 1.48); p = 0.693 |
| ***12 months*** | | | |
| All | 2204 (2030, 2393) [n = 285] | 2208 (2027, 2404) [n = 287] | 1.01 (0.90, 1.14); p = 0.834 |
| ChAdOx1-S | 2270 (1952, 2639) [n = 61] | 1765 (1524, 2045) [n = 61] | 0.78 (0.63, 0.96); p = 0.017 |
| BBIBP-CorV | 2208 (1990, 2449) [n = 192] | 2310 (2072, 2574) [n = 191] | 1.06 (0.92, 1.23); p = 0.434 |
| Gam-COVID-Vac | 2063 (1553, 2740) [n = 32] | 2547 (1957, 3313) [n = 35] | 1.23 (0.86, 1.77); p = 0.256 |

Abbreviations: GM – Geometric Mean. IgG – Immunoglobulin G; GMR – Geometric Mean Ratio; 95% CI – 95 % confidence interval. For baseline, the GMR is adjusted for age group, priming vaccine, duration between first and second dose, duration between second and third (study) dose, and study day of blood draw. For 28 days, 6 and 12 months, the GMR is adjusted for the same variables and baseline anti-spike IgG.

#### *Wuhan-Hu-1 SARS-CoV-2 sVNT inhibition*

Supplementary Table 4 summarises the median percentage inhibition of RBD–hACE2, a surrogate marker for the neutralising capacity against the original SARS-CoV-2 strain (Wuhan-Hu-1) overall and by priming group, stratified by study arm (standard and fractional doses). Baseline and Day 28 results are included for completeness.

*Supplementary Table 4 Median Wuhan-Hu-1 SARS-CoV-2 RBD–hACE2 percentage inhibition by study arm and priming schedule*

| Priming Strata | Median inhibition % (IQR) Standard Dose | Median inhibition % (IQR) Fractional Dose |
|---|---|---|
| **Baseline** | | |
| All | 81 (76–85) [n = 299] | 81 (77–84) [n = 300] |
| ChAdOx1-S | 81 (78–84) [n = 65] | 81 (78–85) [n = 64] |
| BBIBP-CorV | 80 (76–85) [n = 200] | 81 (77–84) [n = 200] |
| Gam-COVID-Vac | 80 (78–84) [n = 34] | 82 (79–84) [n = 36] |
| **Day 28** | | |
| All | 81 (78–84) [n = 292] | 81 (78–84) [n = 295] |
| ChAdOx1-S | 81 (79–84) [n = 65] | 81 (77–84) [n = 62] |
| BBIBP-CorV | 81 (77–84) [n = 194] | 81 (78–83) [n = 198] |
| Gam-COVID-Vac | 80 (76–84) [n = 33] | 81 (79–84) [n = 35] |
| **Six months** | | |
| All | 89 (88–91) [n = 284] | 89 (88–91) [n = 290] |
| ChAdOx1-S | 89 (88–91) [n = 60] | 89 (86–90) [n = 60] |
| BBIBP-CorV | 89 (88–91) [n = 192] | 89 (88–91) [n = 195] |
| Gam-COVID-Vac | 89 (88–91) [n = 32] | 89 (88–91) [n = 35] |
| **12 months** | | |
| All | 89 (88–90) [n = 282] | 89 (87–90) [n = 285] |
| ChAdOx1-S | 89 (88–90) [n = 61] | 89 (88–90) [n = 61] |
| BBIBP-CorV | 89 (88–91) [n = 190] | 89 (87–90) [n = 190] |
| Gam-COVID-Vac | 89 (88–90) [n = 31] | 89 (88–90) [n = 34] |

Abbreviations: IQR – interquartile range

### *Omicron BA.1 SARS-CoV-2 inhibition*

Supplementary Table 5 displays the median RBD–hACE2 percentage inhibition against the SARS-CoV-2 Omicron BA.1 variant at six and twelve months post-booster by priming vaccine and study arm. Baseline and Day 28 results are included for completeness.

*Supplementary Table 5 Median B.1.1.529 SARS-CoV-2 RBD–hACE2 percentage inhibition by study arm and priming schedule*

| Priming Strata | Median inhibition % (IQR) Standard Dose | Median inhibition % (IQR) Fractional Dose |
|---|---|---|
| **Baseline** | | |
| All | 52 (17–77) [n = 299] | 51 (18–76) [n = 300] |
| ChAdOx1-S | 69 (38–81) [n = 65] | 59 (37–75) [n = 64] |
| BBIBP-CorV | 42 (7–72) [n = 200] | 43 (13–72) [n = 200] |
| Gam-COVID-Vac | 61 (36–76) [n = 34] | 68 (41–81) [n = 36] |
| **Day 28** | | |
| All | 82 (75–85) [n = 292] | 81 (75–84) [n = 295] |

| | | |
|---|---|---|
| ChAdOx1-S | 82 (80–84) [n = 65] | 80 (77–83) [n = 62] |
| BBIBP-CorV | 80 (72–85) [n = 194] | 81 (72–84) [n = 198] |
| Gam-COVID-Vac | 83 (78–85) [n = 33] | 82 (76–85) [n = 35] |
| **Six months** | | |
| All | 74 (46–87) [n = 284] | 77 (48–87) [n = 290] |
| ChAdOx1-S | 80 (60–86) [n = 60] | 78 (55–86) [n = 60] |
| BBIBP-CorV | 73 (40–86) [n = 192] | 76 (44–88) [n = 195] |
| Gam-COVID-Vac | 69 (43–88) [n = 32] | 81 (60–88) [n = 35] |
| **12 months** | | |
| All | 76 (44–87) [n = 282] | 79 (40–87) [n = 285] |
| ChAdOx1-S | 82 (65–88) [n = 61] | 74 (50–86) [n = 61] |
| BBIBP-CorV | 72 (35–87) [n = 190] | 79 (37–87) [n = 190] |
| Gam-COVID-Vac | 79 (49–88) [n = 31] | 84 (62–89) [n = 34] |

Abbreviations: IQR – interquartile range

#### *Documented SARS-CoV-2 infections*

Documented SARS-CoV-2 infections are shown in Supplementary Figure 1 (n = 25).

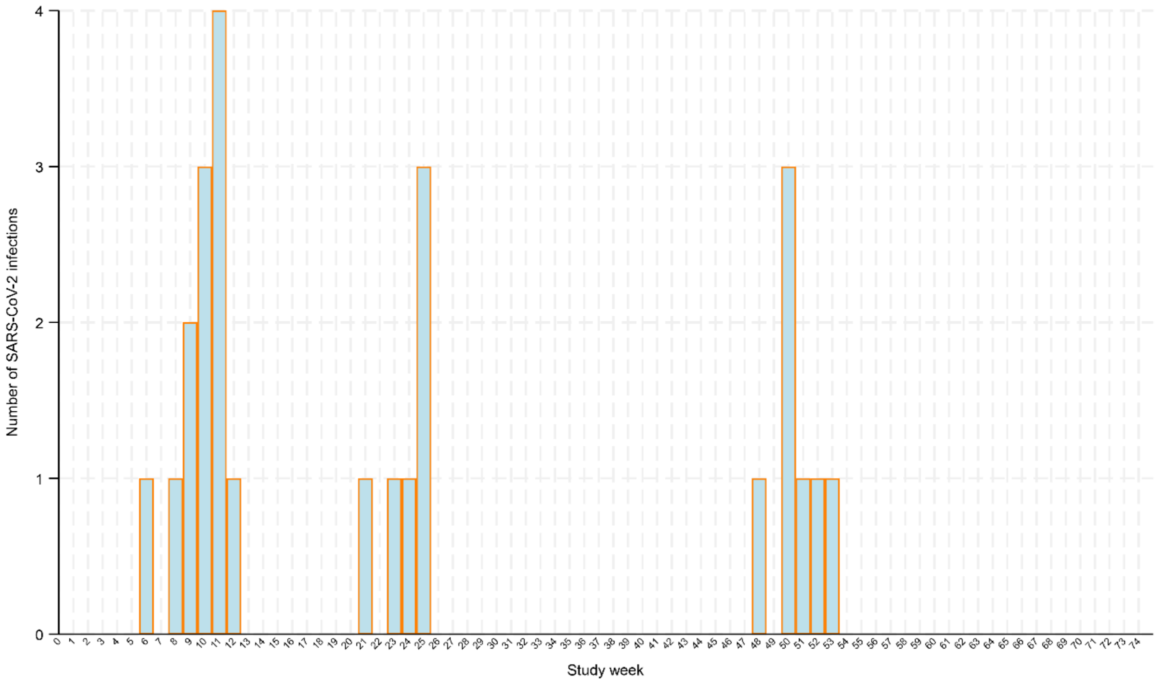

*Supplementary Figure 1 SARS-CoV-2 infections by study week from trial baseline to 12 month visit window (n = 25)*

### Adverse and serious adverse events

#### *Adverse events*

Supplementary Table 6 provides an overview of adverse events (AEs) reported among participants, classified by their relationship to the study vaccine, severity, and timelines.

*Supplementary Table 6 Adverse events (n=76), classified by study arm, MedDRA term, relationship to study vaccine, severity, onset relative to vaccination, duration, and outcome*

| Case number | Study arm | MedDRA term | Relationship to Study Vaccine [a] | Severity [b] | Administration of study vaccine to AE start date (days) | Duration of event (days) | Outcome |
|---|---|---|---|---|---|---|---|
| 1 | Standard | Abdominal cramps | Unrelated | Moderate | 3 | 0 | Resolved |
| 2 | Standard | Arthralgia | Unrelated | Moderate | 1 | 40 | Resolved |
| 3 | Standard | Back pain | Unrelated | Mild | 175 | 2 | Resolved |
| 4 | Standard | Back pain | Unrelated | Mild | 21 | 56 | Resolved |
| 5 | Standard | Blood glucose increased | Unrelated | Mild | 55 | 0 | Resolved |
| 6 | Standard | Chronic bronchitis | Unrelated | Mild | 173 | 40 | Resolved |
| 7 | Standard | Dyspnoea | Unrelated | Mild | 6 | 20 | Resolved with sequelae |
| 8 | Standard | Eczema | Possible | Mild | 17 | 26 | Resolved |
| 9 | Standard | Eye inflammation | Unrelated | Mild | 64 | 6 | Resolved |
| 10 | Standard | Fatigue | Possible | Mild | 2 | 24 | Resolved |
| 11 | Standard | Furunculosis | Possible | Moderate | 33 | 62 | Resolved |
| 12 | Standard | Gallbladder disorder | Unrelated | Mild | 94 | 14 | Resolved |
| 13 | Standard | Gallstones | Unrelated | Mild | 174 | 7 | Resolved |
| 14 | Standard | Headache | Possible | Mild | 16 | 69 | Resolved |
| 15 | Standard | Headache | Possible | Mild | 0 | 27 | Resolved |
| 16 | Standard | Headache | Possible | Mild | 8 | 18 | Resolved |
| 17 | Standard | Headache | Possible | Moderate | 8 | 14 | Resolved |
| 18 | Standard | Headache | Possible | Moderate | 48 | 12 | Resolved |
| 19 | Standard | Headache | Unrelated | Moderate | 0 | 37 | Resolved with sequelae |
| 20 | Standard | Hypertension | Unrelated | Mild | 25 | 3 | Resolved |
| 21 | Standard | Hypertension | Unrelated | Mild | 328 | 8 | Resolved |
| 22 | Standard | Hypertension | Possible | Mild | 7 | 8 | Resolved |
| 23 | Standard | Hypertension | Possible | Moderate | 83 | 10 | Resolved with sequelae |
| 24 | Standard | Hypertension | Possible | Moderate | 10 | 9 | Resolved with sequelae |
| 25 | Standard | Menstruation irregular | Possible | Moderate | 3 | 99 | Resolved |
| 26 | Standard | Natural menopause | Possible | Mild | 0 | 109 | Resolved |
| 27 | Standard | Radius fracture | Unrelated | Mild | 190 | 5 | Resolved |

| | | | | | | | |
|---|---|---|---|---|---|---|---|
| 28 | Standard | Upper respiratory tract infection | Unrelated | Mild | 26 | 7 | Resolved |
| 29 | Standard | Upper respiratory tract infection | Unrelated | Moderate | 1 | 6 | Resolved |
| 30 | Standard | Dental care | Unrelated | Mild | 56 | 21 | Resolved |
| 31 | Standard | Dental care | Unrelated | Mild | 69 | 0 | Resolved |
| 32 | Standard | Dental care | Unrelated | Mild | 84 | 2 | Resolved |
| 33 | Standard | Dental care | Unrelated | Mild | 91 | 8 | Resolved |
| 34 | Standard | Eye injury | Unrelated | Moderate | 76 | 11 | Resolved |
| 35 | Standard | Limb injury | Unrelated | Mild | 106 | 21 | Resolved |
| 36 | Standard | Acute upper respiratory tract infection | Unrelated | Mild | 22 | 5 | Resolved |
| 37 | Standard | Acute respiratory tract infection | Unrelated | Moderate | 10 | 22 | Resolved |
| 38 | Standard | Blood loss anaemia | Unrelated | Moderate | 65 | 134 | Resolved |
| 39 | Fractional | Amenorrhea | Possible | Mild | 45 | 70 | Resolved |
| 40 | Fractional | Anaemia | Unrelated | Moderate | 20 | 7 | Resolved |
| 41 | Fractional | Arthralgia | Unrelated | Moderate | 1 | 10 | Resolved |
| 42 | Fractional | Back injury | Unrelated | Moderate | 132 | 7 | Resolved with sequelae |
| 43 | Fractional | Back pain | Unrelated | Moderate | 8 | 12 | Resolved |
| 44 | Fractional | Chest discomfort | Unrelated | Mild | 11 | 16 | Resolved with sequelae |
| 45 | Fractional | Chest pain | Possible | Moderate | 44 | 5 | Resolved |
| 46 | Fractional | Chronic gastritis | Unrelated | Mild | 56 | 14 | Resolved |
| 47 | Fractional | Dental caries | Unrelated | Mild | 144 | 3 | Resolved |
| 48 | Fractional | Diabetes mellitus | Unrelated | Mild | 79 | 0 | Resolved |
| 49 | Fractional | Furuncle | Possible | Mild | 12 | 8 | Resolved |
| 50 | Fractional | Furuncle | Possible | Mild | 27 | 8 | Resolved |
| 51 | Fractional | Headache | Possible | Mild | 1 | 30 | Resolved |
| 52 | Fractional | Headache | Possible | Mild | 23 | 4 | Resolved |
| 53 | Fractional | Headache | Possible | Mild | 8 | 6 | Resolved |
| 54 | Fractional | Headaches | Unrelated | Moderate | 143 | 31 | Resolved |
| 55 | Fractional | Heart rate irregular | Possible | Mild | 31 | 84 | Resolved with sequelae |
| 56 | Fractional | Hypersensitivity | Unrelated | Moderate | 67 | 3 | Resolved |
| 57 | Fractional | Hypertension | Possible | Mild | 8 | 97 | Resolved |
| 58 | Fractional | Hypertension | Possible | Mild | 39 | 45 | Resolved with sequelae |
| 59 | Fractional | Hypertension | Unrelated | Moderate | 8 | 7 | Resolved |
| 60 | Fractional | Irregular menstruation | Possible | Mild | 8 | 109 | Resolved |

| | | | | | | | |
|---|---|---|---|---|---|---|---|
| 61 | Fractional | Menstruation irregular | Possible | Moderate | 39 | 75 | Resolved |
| 62 | Fractional | Myalgia | Unrelated | Mild | 64 | 44 | Resolved |
| 63 | Fractional | Palpitations | Possible | Moderate | 3 | 12 | Resolved |
| 64 | Fractional | Pregnancy | Unrelated | Mild | 116 | 61 | Resolved |
| 65 | Fractional | Soft tissue injury | Unrelated | Mild | 163 | 15 | Resolved |
| 66 | Fractional | Syncope | Unrelated | Mild | 37 | 0 | Resolved |
| 67 | Fractional | Upper respiratory tract infection | Unrelated | Moderate | 2 | 27 | Resolved |
| 68 | Fractional | Varicella | Unrelated | Moderate | 344 | 7 | Resolved |
| 69 | Fractional | Tooth infection | Unrelated | Moderate | 16 | 7 | Resolved |
| 70 | Fractional | Face injury | Unrelated | Mild | 86 | 8 | Resolved |
| 71 | Fractional | Lower limb fracture | Unrelated | Mild | 103 | 20 | Resolved |
| 72 | Fractional | Allergy to chemicals | Unrelated | Moderate | 24 | 5 | Resolved |
| 73 | Fractional | Acute upper respiratory tract infection | Unrelated | Mild | 21 | 5 | Resolved |
| 74 | Fractional | Acute respiratory tract infection | Unrelated | Mild | 23 | 5 | Resolved |
| 75 | Fractional | Acute respiratory tract infection | Unrelated | Moderate | 18 | 6 | Resolved |
| 76 | Fractional | Dust allergy | Possible | Mild | 70 | 36 | Resolved |

**Abbreviations:** AE – adverse event; MedDRA – Medical Dictionary for Regulatory Activities. **Footnotes:** [a] Relationship to study vaccine was categorised as unrelated, possible, probable, or definite. [b] Severity was graded as mild (grade 1), moderate (grade 2), severe (grade 3), potentially life-threatening (grade 4), or fatal (grade 5).

#### *Serious adverse events*

Supplementary Table 7 shows all serious adverse events (SAEs) recorded within the 12 month visit window.

*Supplementary Table 7 Serious adverse events (n=41), classified by study arm, MedDRA term, relationship to study vaccine, severity, onset relative to vaccination, duration, and outcome*

| **Case number** | **Study arm** | **MedDRA term** | **Relationship to Study vaccine [a]** | **Severity [b]** | **Administration of study vaccine to AE start date (days)** | **Duration of event (days)** | **Outcome** |
|---|---|---|---|---|---|---|---|
| 1 | Standard | Back pain | Unrelated | Moderate | 38 | 8 | Resolved |
| 2 | Standard | Cholecystectomy | Unrelated | Severe | 118 | 5 | Resolved |
| 3 | Standard | Cholecystectomy | Unrelated | Severe | 88 | 3 | Resolved |
| 4 | Standard | Cholecystitis | Unrelated | Potentially life-threatening | 11 | 11 | Resolved with sequelae |
| 5 | Standard | Chronic bronchitis | Unrelated | Severe | 35 | 7 | Resolved with sequelae |
| 6 | Standard | Colorectal cancer stage II | Unrelated | Severe | 293 | 29 | Resolved with sequelae |
| 7 | Standard | Cystocele | Unrelated | Moderate | 177 | 7 | Resolved |
| 8 | Standard | Diabetes mellitus | Unrelated | Moderate | 294 | 7 | Resolved with sequelae |
| 9 | Standard | Diabetes mellitus | Unrelated | Severe | 17 | 36 | Resolved with sequelae |
| 10 | Standard | Diabetes mellitus | Unrelated | Severe | 82 | 6 | Resolved with sequelae |

| | | | | | | | |
|---|---|---|---|---|---|---|---|
| 11 | Standard | Diabetes mellitus inadequate control | Unrelated | Severe | 45 | 10 | Resolved with sequelae |
| 12 | Standard | Ectopic pregnancy | Unrelated | Severe | 130 | 5 | Resolved |
| 13 | Standard | Gastric cancer | Unrelated | Fatal | 44 | 87 | Fatal† |
| 14 | Standard | Hypertension | Unrelated | Moderate | 46 | 7 | Resolved with sequelae |
| 15 | Standard | Pyelonephritis chronic | Unrelated | Moderate | 385 | 7 | Resolved |
| 16 | Standard | Respiratory infection | Unrelated | Moderate | 99 | 7 | Resolved |
| 17 | Standard | Lower limb fracture | Unrelated | Severe | 1 | 13 | Resolved |
| 18 | Standard | Lower limb fracture | Unrelated | Severe | 64 | 273 | Resolved |
| 19 | Standard | Meniscus injury | Unrelated | Severe | 369 | 148 | Resolved |
| 20 | Standard | Decompensated diabetes | Unrelated | Fatal | 124 | 40 | Fatal† |
| 21 | Fractional | Arm amputation | Unrelated | Potentially life-threatening | 352 | 19 | Resolved with sequelae |
| 22 | Fractional | Asthma, unspecified type, with status asthmaticus | Unrelated | Severe | 154 | 6 | Resolved with sequelae |
| 23 | Fractional | Back injury | Unrelated | Severe | 84 | 5 | Resolved with sequelae |
| 24 | Fractional | Back pain | Unrelated | Moderate | 78 | 13 | Resolved |
| 25 | Fractional | Back pain | Unrelated | Severe | 160 | 7 | Resolved with sequelae |
| 26 | Fractional | Cerebrovascular accident | Unrelated | Severe | 322 | 11 | Resolved with sequelae |
| 27 | Fractional | Cholecystitis | Unrelated | Severe | 21 | 93 | Resolved |
| 28 | Fractional | Cholelithiasis | Unrelated | Moderate | 139 | 31 | Resolved |
| 29 | Fractional | Chronic pyelonephritis | Unrelated | Moderate | 265 | 7 | Resolved |
| 30 | Fractional | Completed suicide | Unrelated | Fatal | 263 | 0 | Fatal† |
| 31 | Fractional | Diabetes mellitus | Unrelated | Severe | 98 | 7 | Resolved with sequelae |
| 32 | Fractional | Gallbladder disorder | Unrelated | Moderate | 374 | 68 | Resolved |
| 33 | Fractional | Gout | Unrelated | Moderate | 11 | 21 | Resolved with sequelae |
| 34 | Fractional | Gouty arthritis | Unrelated | Severe | 51 | 11 | Resolved with sequelae |
| 35 | Fractional | Haemorrhoids | Unrelated | Severe | 323 | 5 | Resolved |
| 36 | Fractional | Headache | Unrelated | Moderate | 126 | 7 | Resolved with sequelae |
| 37 | Fractional | Hypertension | Unrelated | Moderate | 80 | 7 | Resolved |
| 38 | Fractional | Hypertension | Unrelated | Moderate | 292 | 10 | Resolved with sequelae |
| 39 | Fractional | Pneumonia mycoplasmal | Unrelated | Severe | 166 | 18 | Resolved |
| 40 | Fractional | Cerebral cyst | Unrelated | Moderate | 246 | 167 | Resolved |
| 41 | Fractional | Ulcerative gastritis | Unrelated | Severe | 153 | 7 | Resolved with sequelae |

**Abbreviations:** AE – adverse event; MedDRA – Medical Dictionary for Regulatory Activities. **Footnotes:** [a] Relationship to study vaccine was categorised as unrelated, possible, probable, or definite. [b] Severity was graded as mild (grade 1), moderate (grade 2), severe (grade 3), potentially life-threatening (grade 4), or fatal (grade 5).
